# Preliminary outcomes of combined treadmill and overground high-intensity interval training in ambulatory chronic stroke

**DOI:** 10.1101/2021.10.25.21265471

**Authors:** Pierce Boyne, Sarah Doren, Victoria Scholl, Emily Staggs, Dustyn Whitesel, Daniel Carl, Rhonna Shatz, Russel Sawyer, Oluwole O. Awosika, Darcy S. Reisman, Sandra A. Billinger, Brett Kissela, Jennifer Vannest, Kari Dunning

**Author notes:** **Corresponding author:** Pierce Boyne, PT, DPT, PhD, NCS, Health Sciences Building, room 267, 3225 Eden Ave., Cincinnati, OH, 45267-0394.

## Abstract

**Introduction:** Locomotor high-intensity interval training (HIIT) is a promising intervention for stroke rehabilitation that typically involves bursts of fast treadmill walking alternated with recovery periods. However, overground translation of treadmill speed gains has been somewhat limited, some important outcomes have not been tested and baseline response predictors are poorly understood. This pilot study aimed to guide future research by assessing preliminary outcomes of combined overground and treadmill HIIT.

**Methods:** Ten participants >6 months post stroke completed a multi-domain assessment battery before and after a 4-week no-intervention control phase, then again after a 4-week treatment phase involving 12 sessions of overground and treadmill HIIT. The primary analyses assessed relative changes in overground and treadmill walking speeds after HIIT, evaluated responsiveness of different outcome measures and estimated effects of baseline gait speed on treatment response.

**Results:** Overground and treadmill gait function both improved during the treatment phase relative to the control phase, with overground speed changes averaging 61% of treadmill speed changes (95% CI: 33-89%). Moderate or larger effect sizes were observed for measures of gait performance, balance, fitness, cognition, fatigue, perceived change and brain volume. Participants with baseline comfortable gait speed <0.4 m/s had less absolute improvement in walking capacity but similar proportional and perceived changes.

**Discussion:** Future locomotor HIIT studies should consider including: 1) both overground and treadmill training; 2) measures of cognition, fatigue and brain volume, to complement typical motor & fitness assessment; and 3) baseline gait speed as a covariate.

## INTRODUCTION

Locomotor high-intensity interval training (HIIT) is a promising new intervention for stroke rehabilitation.^1,2^ This method involves bursts of fast walking alternated with recovery periods to enable vigorous aerobic intensities, using burst and recovery intervals ranging from short (e.g. 30 seconds) to long (e.g. 3-4 minutes).^3^ Initial longitudinal stroke studies suggest that both short-interval and long-interval locomotor HIIT are feasible, can improve gait function & exercise capacity,^4-11^ and could be more efficacious than conventional moderate-intensity exercise.^6,7^

However, locomotor HIIT has only been tested on a treadmill, and treadmill-only exercise often has limited translation to overground gait improvements after stroke.^6,12,13^ Based on the neuroplasticity principle of task-specificity,^14^ we proposed combining overground and treadmill HIIT to improve overground translation.^6,15 14^This method showed feasibility in ambulatory chronic stroke,^15^ but no previous studies have reported outcome changes after a protocol involving overground HIIT. Such information could help determine whether further protocol refinement may be needed before considering an efficacy trial.

Another limitation to prior post-stroke HIIT studies is that they have mostly included only gait & exercise capacity outcomes. HIIT also has the theoretical potential to facilitate other important aspects stroke recovery, such as cognition, fatigue and brain health.^16-18^ In addition, there is reason to suspect that some of these potential HIIT effects could be greater than those elicited by moderate intensity exercise.^16,17^ For example, HIIT has been shown to elicit greater acute increases in some circulating neurotrophins than moderate intensity exercise after stroke.^19,20^ However, no longitudinal stroke studies have assessed changes in cognition, fatigue or brain volumes after locomotor HIIT. Thus, it remains unknown which outcome measures, if any, might be sufficiently responsive to consider including in future studies.

Additionally, there is limited available information about baseline patient characteristics associated with responsiveness to HIIT post-stroke. Previous studies have shown that stroke survivors with slower baseline walking speed tend to have less absolute improvement in gait function with various forms of training.^21-24^ Yet, it is uncertain if this generalizes to HIIT and whether proportional change or perception of improvement are similar between participants with different baseline walking speeds. This knowledge could help guide selection of eligibility criteria, stratification factors and covariates in future studies.

The aims of this pilot study were to: 1) Assess overground translation of treadmill speed gains after combined overground and treadmill HIIT in chronic hemiparetic stroke; 2) Evaluate responsiveness of different outcome measures across various domains, including gait, balance, exercise capacity, cognitive, fatigue/subjective and brain volume measures, to guide selection of outcome measures for future trials; and 3) Examine the potential for differences in responsiveness between participants with faster vs. slower baseline gait speed. Participants >6 months post-stroke had a no-intervention control phase followed by 12 sessions of combined overground and treadmill HIIT (6 sessions of short-interval HIIT alternated with 6 sessions of long-interval HIIT). We hypothesized that: 1) On average, at least 50% of treadmill speed gains during the treatment phase would translate into overground gait; 2) Standardized effect size estimates in each outcome domain would be ≥0.4 between phases, justifying further study;^25^ and 3) Compared with participants who have baseline comfortable gait speed ≥0.4 m/s, those with speed <0.4 m/s would have less absolute improvement in 6-minute walk distance,^21^ but similar proportional change and perception of improvement.

## METHODS

The study providing data for this analysis was approved by institutional review boards, preregistered on ClinicalTrials.gov" (NCT02858349) and performed in a cardiovascular stress laboratory, MRI research center and rehabilitation research laboratory from July 2016 to December 2017. Some of the methods and baseline data from this study have been previously described in manuscripts addressing different aims from the current report.^15,26^

### Participants

Participants with ischemic or hemorrhagic stroke and control participants without stroke were recruited from the community and provided written informed consent. To be eligible, participants had to be 30-90 years old, MRI compatible, able to communicate with investigators and correctly answer consent comprehension questions, able to perform mental imagery,^27^ without recent history of drug/alcohol abuse or significant mental illness, and not pregnant. Participants with stroke had to meet the following additional criteria: unilateral stroke in middle cerebral artery territory experienced >6 months prior to enrollment; walking speed <1.0 m/s on the 10 meter walk test^28^; able to walk 10m over ground with assistive devices as needed and no physical assistance; no evidence of significant arrhythmia or myocardial ischemia on treadmill ECG stress test, or significant baseline ECG abnormalities that would make an exercise ECG uninterpretable^29^; no recent cardiopulmonary hospitalization; no significant ataxia or neglect (NIHSS item score >1)^30^; no severe lower extremity (LE) hypertonia (Ashworth >2)^31^; no major post-stroke depression (PHQ-9 ≥10)^32^ in the absence of management of the depression by a health care provider^33^; not participating in physical therapy or another interventional research study; no recent paretic LE botulinum toxin injection; and no concurrent progressive neurologic disorder or other major conditions that would limit capacity for improvement. Control participants had to be a demographic match for a participant with stroke (same sex, age difference ≤5 years) without any current neurologic, orthopedic or medical condition affecting gait.

### Study design

After baseline assessment, each participant with stroke had a 4-week control phase with no-intervention, followed by a 4-week treatment phase with 12 sessions of locomotor HIIT, approximately 3x/week. In alternating order, 6 of these sessions used short-interval HIIT while the other 6 used long-interval HIIT, with the first session randomized across participants. Outcome measures were assessed before the control phase (PRE), after the control phase (4WK), and after the 4-week treatment phase (8WK). Control participants had baseline testing (PRE) only.

### Screening

Eligibility determination was based on medical history review and physical assessment. Participants with stroke also had treadmill acclimation and graded exercise testing (GXT) with electrocardiography for screening, as previously described.^6,34^ For the GXT, treadmill speed was held constant at a pre-determined individualized value, while incline was increased 2-4% until volitional fatigue, severe gait instability or a cardiovascular safety limit.^29,35^

### Outcome testing

The following outcome measures were assessed for participants with stroke at PRE, 4WK and 8WK by a blinded physical therapist who had not been involved with the participant’s treatment. For the 4WK and 8WK testing, the testing therapist did not know whether the participant had received any treatment in the previous phase. Participants used any habitual orthotic and assistive devices during gait testing and were guarded as needed, but only assisted if required to prevent a fall or injury. These measures were assessed for control participants one time only (PRE).

#### Gait function

- The 10-meter walk test^36^ was performed twice at comfortable speed and twice at fastest speed with trial pairs averaged for analysis to calculate comfortable and fastest overground gait speeds.
- The 6-minute walk test was performed and total distance was recorded.^37^

#### Spatiotemporal gait parameters

- Two passes were performed at comfortable speed across a sensor-embedded electronic walkway^38,39^ and were combined for analysis, with parameters averaged across gait cycles, including: cadence, step time symmetry, step lengths & symmetry and single-limb support times & symmetry. Symmetry indices were calculated as: (1 - | Paretic – Nonparetic | / (Paretic + Nonparetic)) * 100%; with possible values ranging from 0-100%, where 0% means complete asymmetry and 100% means perfect symmetry.

The following outcome measures were assessed for participants with stroke by the treating physical therapist and a research assistant (both unblinded), on a separate day from the gait measures above, at each testing time point. Control participants were also assessed on these measures at PRE except as indicated below.

#### Gait function

- Fastest safe treadmill speed was evaluated with a rapid acceleration test, where speed was increased by 0.1 mph every 5 seconds until the participant could no longer maintain the speed or had severe gait instability. This test was not done for controls.

#### Standing balance

- The NIH Toolbox Standing Balance Test Age 7+^40^ was performed using the NIH Toolbox iPad application and BalancePod iPod touch application with recommended equipment.^40^ Fully corrected T scores were used for analysis. These scores are referenced to persons with the same age, gender, race/ethnicity and educational attainment as the participant from the NIH Toolbox nationally representative normative sample,^41-43^ with a mean of 50 and a standard deviation of 10. Thus, a score of 40 would represent worse balance than the average American with similar demographics, one standard deviation below the mean.

#### Exercise capacity

These assessments were done only for participants with stroke. A GXT was performed following the same protocol as the screening GXT, but with a metabolic cart and facemask for gas exchange analysis.^44^ This was done on a separate day from the screening GXT at PRE. Fifteen minutes after the GXT, participants performed another exercise test for verification of peak exercise capacity. This test started at the participant’s pre-determined fastest safe speed and 2-4% grade. Incremental grade then speed decreases were done as little as needed to enable continued walking as fast as possible for 3 minutes, with close guarding.

- The oxygen consumption rate (VO_2_) at the ventilatory threshold during the GXT was determined using a combination of the V-slope and ventilatory equivalents methods, as previously described.^45^
- The highest VO_2_ and heart rate (HR) values from either the GXT or 3-minute test were recorded as VO_2peak_ and HR_peak_.

#### Cognition

- The Free and Cued Selective Reminding Test (FCSRT)^46,47^ was administered and the free recall score was used to assess episodic memory. The FCSRT was found to be the most accurate clinical test for identifying prodromal Alzheimer disease among patients with mild cognitive impairment and had an optimal free recall cutoff score of <17 (on a scale from 0-48).^48^ This test was not done for controls.
- The following tests from the NIH Toolbox Cognition Battery^49^ were administered using the NIH Toolbox iPad application: Flanker inhibitory control & attention test (assesses attention and executive functioning), picture sequence memory test (episodic memory), list sorting working memory test (working memory), dimensional change card sort test (executive function) and pattern comparison processing speed test (processing speed). The NIH Toolbox also provides a fluid cognition composite score combining the results from each of these tests. Fully corrected T scores were used for analysis.

#### Subjective measures

These assessments were done only for participants with stroke.

- The Patient Reported Outcomes Measurement Information System (PROMIS^®^) Fatigue^50,51^ short form 8a v1.0 (PROMIS-Fatigue) was administered, and raw scores were converted to T scores according to the scoring manual. These T scores are referenced to the entire U.S. general population PROMIS calibration sample,^52^ with a mean of 50 and a standard deviation of 10. Thus, a score of 60 would represent greater fatigue than the average American, one standard deviation above the mean.
- The Stroke and Aphasia Quality of Life Scale 39^53^ was administered and a total score from 1-5 was calculated by averaging all items, where a score of 1 on each item strongly endorses (definitely yes) and a score of 5 strongly denies (definitely no) a particular problem or difficulty.
- Global Ratings of Change (GROC)^54^ for overall stroke recovery, walking ability and fitness were administered at 4WK and 8WK (rating *change* is not applicable at PRE). Participants were asked to rate the change in each characteristic from the time that they began the study until that testing session, using a 15-point scale ranging from -7 (a very great deal worse) to 0 (about the same) to +7 (a very great deal better).^54^ Scores of +1 (hardly any better at all) to +3 (somewhat better) are considered to represent small improvement, while scores of +4 (moderately better) to +5 (a good deal better) represent moderate improvement and scores of +6 (a great deal better) and +7 represent large improvement.^54^

#### Brain volume

Structural MRI acquisition and preprocessing has been previously described in detail.^26^ Briefly, T1-weighted (T1w) brain images were obtained with a 3.0T Philips Ingenia MRI system (1 mm isotropic resolution; TR, 8.1 ms; TE, 3.7 ms; flip angle, 8^0^; SENSE factor 2). FSL software^55^ was used for bias field correction, tissue type segmentation and non-linear registration to the MNI152 template, using a lesion mask to improve registration for participants with stroke. Lesioned voxels in the native-space T1w image were temporarily filled with MNI template voxels^56^ to enable automated structure labelling with FreeSurfer software.^57^ Lesioned voxels were then masked out of any structure labels.

- Supratentorial brain volume (excluding brainstem and cerebellum) was calculated bilaterally and for the ipsilesional & contralesional hemibrain by summing the number of voxels in gray and white matter structure labels and multiplying by the voxel size. These volumes were adjusted for head size by first calculating the scaling factor from the participant’s skull to the MNI152 template skull (larger scaling factor = smaller participant skull),^58^ then effectively regressing the scaling factor out of the volumetric data.^59,60^ This adjustment used the formula:

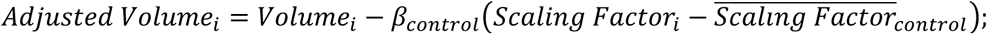

where the subscript “i” represents values from the individual participant scan, β_control_ is the slope of the regression line from using the scaling factor to predict the volume of interest in the control group, and 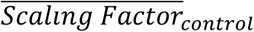 is the mean scaling factor in the control group.^60^ For bilateral, ipsilesional and contralesional (supratentorial) brain volumes, β_control_ values were -712.7, -358.4 and -354.3, respectively (all p<0.0001). 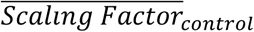 was 1.27.

### Locomotor HIIT treatment protocol

The treatment protocol and training intensity data have been previously reported in detail.^15^ Briefly, each session included a 3-minute warm up (overground walking at ∼40% HR reserve [HRR]), 10 minutes of overground HIIT, then 20 minutes of treadmill HIIT, followed by another 10 minutes of overground HIIT and a 2-minute cool down (overground walking at ∼40% HRR). HRR bpm targets were calculated by: (HR_peak_ – HR_resting_) x %HRR target + HR_resting_.^29^ Participants wore habitual orthotic devices, started with habitual assistive devices during overground training and used a fall protection harness during treadmill training. Physical assistance was only provided if needed to avoid a fall or injury. During overground training, participants walked back and forth in a corridor, using visual feedback about distance covered and verbal encouragement to maximize speed each burst. Assistive device and/or pattern were progressed if it enabled achievement of faster speeds.

Each participant alternated between short and long-interval HIIT sessions. Short-interval HIIT sessions involved 30 second bursts at maximum safe speed alternated with 30-60 second resting recovery periods.^6^ Burst speed was progressed as able throughout the session to maintain constant challenge. Long interval HIIT sessions involved 3-4 minute bursts at a target HR of 90% HR_peak_ alternated with 2-3 minute active recovery periods at a target of 70% HR_peak_. Speed was continually adjusted as needed to achieve and maintain the target HR.

During short-interval HIIT sessions, actual mean training speed was 0.75 m/s overground and 0.90 m/s on the treadmill, while mean training HR was 78% and 83% HR_peak_ respectively.^15^ During long-interval HIIT sessions, mean speed was 0.67 m/s overground and 0.51 m/s on the treadmill, while mean training HR was 81% and 82% HR_peak_ respectively.^15^

### Safety monitoring

Adverse events (AEs) were defined as any undesirable change in participant health (regardless of severity or suspected relationship with the study procedures) and were systematically assessed. Participants with stroke were asked about AEs at each visit, were specifically queried about soreness/pain, fatigue, nausea, lightheadedness, falls and injuries, and were continuously monitored for AEs throughout each testing and treatment session.

### Data analysis

Baseline measures were compared between stroke survivors and controls using independent t-tests and Fisher exact tests. Baseline values were also expressed as a percentage of normative demographic-predicted values for comfortable gait speed,^28^ 6-minute walk test,^61^ HR_peak_^62,63^ and VO_2peak_. ^64^ AEs were categorized by relationship to the intervention, type and severity.^65^ To estimate group-level outcome changes within and between phases for the stroke group, mixed effects general linear models were used with the outcome of interest (e.g. comfortable gait speed) as the dependent variable, a fixed (categorical) effect for time (PRE, 4WK, 8WK) and an unstructured covariance matrix to account for repeated measures within the same participant. SAS version 9.4 was used for analysis.

For hypothesis 1 (at least 50% of treadmill speed gains would translate into overground gait), we calculated change in fastest 10-meter walk test speed during the treatment phase, expressed it as a percentage of change in fastest treadmill speed and averaged across participants.

For hypothesis 2 (standardized effect sizes in each outcome domain would be ≥0.4), we calculated Cohen’s d effect sizes^66^ for each outcome measure during the treatment phase by dividing the mean change by the standard deviation of change and multiplying by √2. We also performed the same calculation for the treatment vs. control phase comparison using individual values for treatment phase change minus control phase change.

For hypothesis 3 (faster walkers at baseline would have greater absolute improvement in 6-minute walk distance, but similar proportional change and perception of improvement), we used the mixed effects model described above and added fixed effects for baseline comfortable gait speed subgroup (< 0.4 m/s vs. ≥ 0.4 m/s)^21^ and its interaction with time. Preliminary subgroup estimates were calculated for treatment phase changes in gait function (absolute and percent changes) and subjective measures.

### Target sample size

Based on previous study aims,^26^ the target sample size was 10 stroke survivors and 10 controls with complete data. At the 0.05 significance level, this sample size provides 80% power to detect a within-group change Cohen’s d effect size as small as 1.00 (very large) and a between group (stroke vs. control) Cohen’s d effect size as small as 1.32. Therefore, this pilot study was only powered to statistically detect very large or greater effects.

## RESULTS

Ten participants with stroke and 10 healthy controls were enrolled (Fig 1). All participants completed the trial, with no missing visits. Compared with controls, participants with stroke were significantly less independent with ambulation, used more orthotic & assistive devices and had lower gait speed & 6-minute walk distance (Table 1). Participants with stroke also had significantly lower values for various spatiotemporal gait parameters, standing balance, fluid cognition and brain volume (Table 2). No serious AEs occurred throughout the study. Mild and moderate AEs are reported in Table 3.

**Figure 1.**
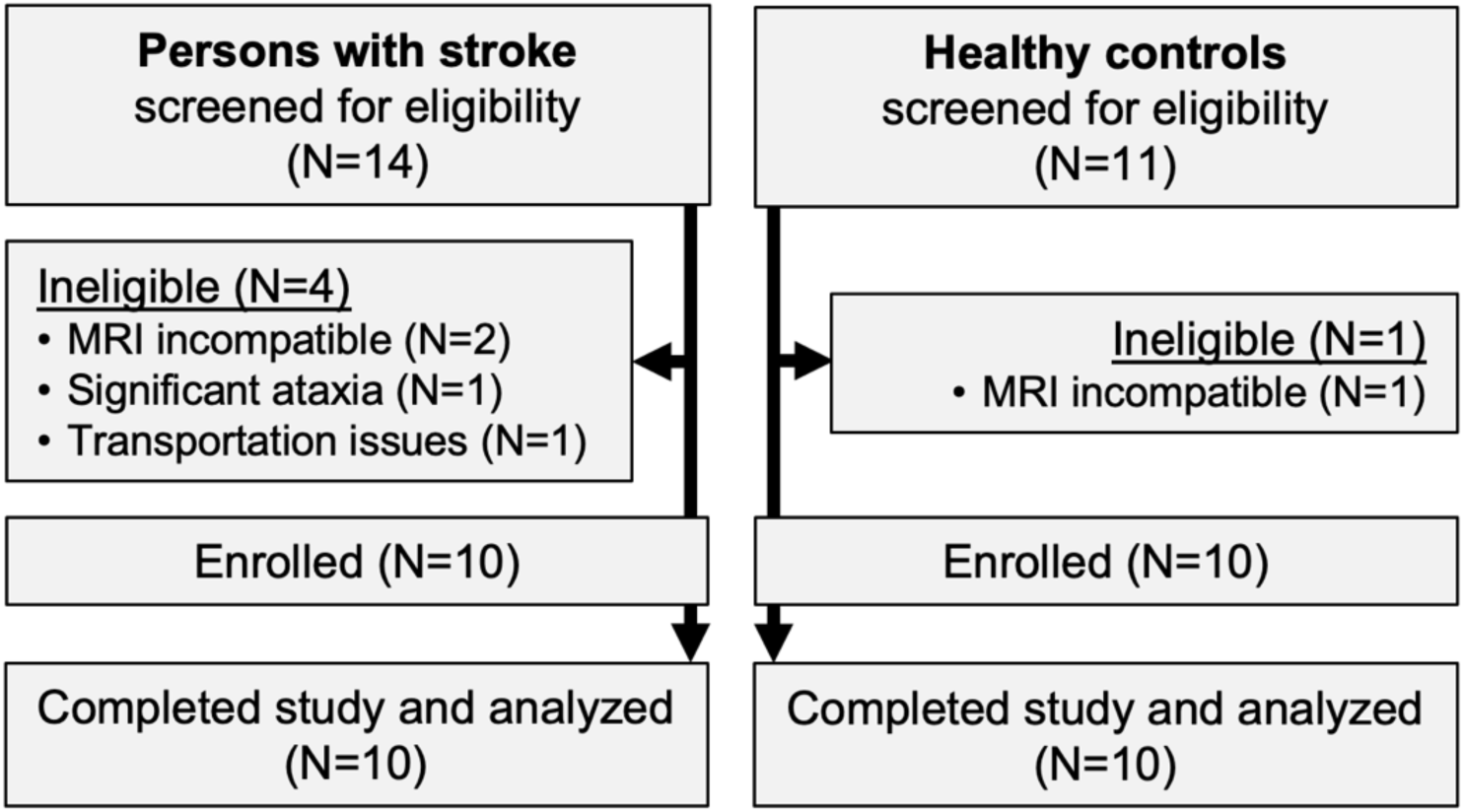
Study flow diagram.

**Table 1.**
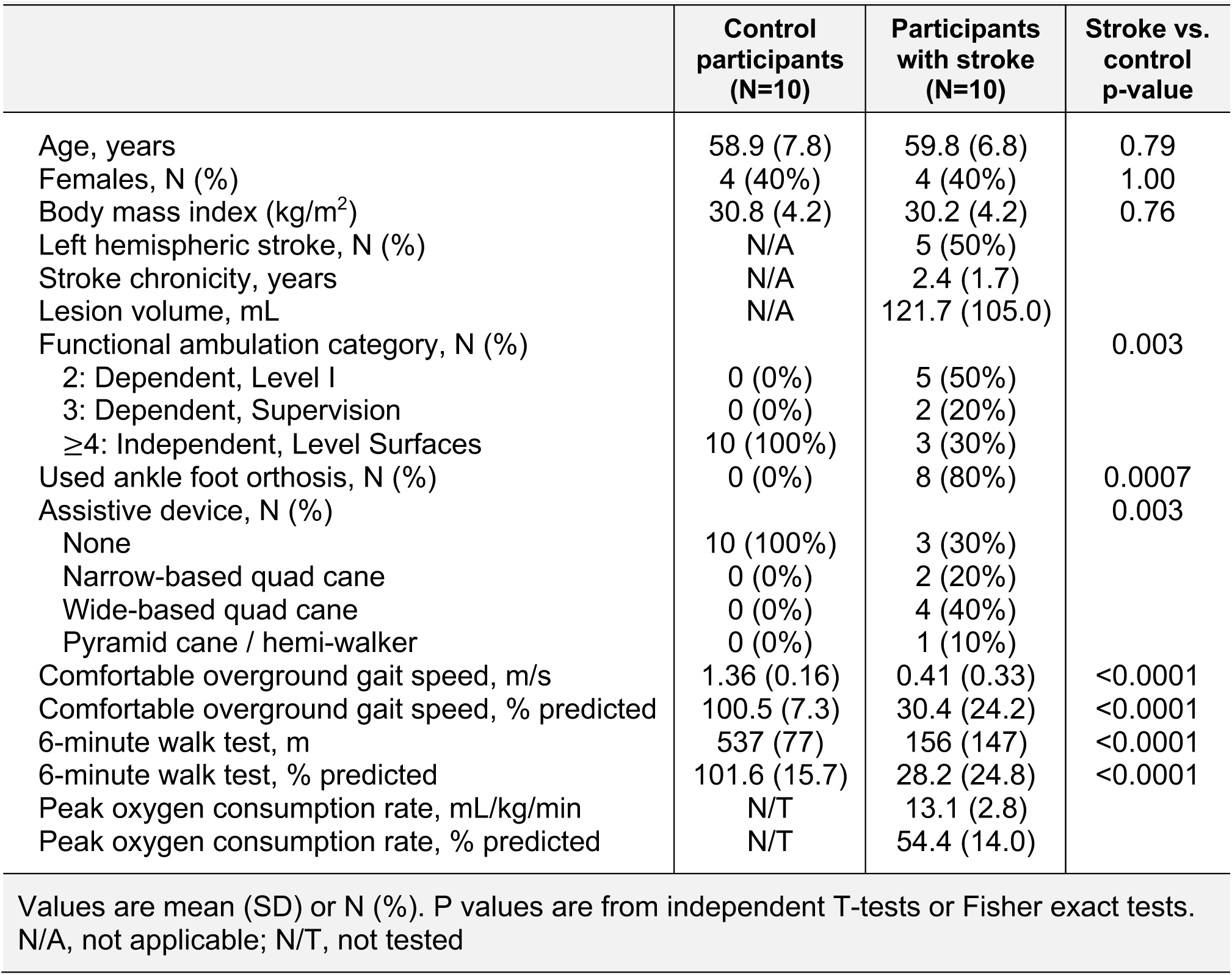
Participant characteristics.

**Table 2.** Adverse events (AEs) among participants with stroke (N=10; total visits=200)*

|  | No. (%) of participants with AE | Total number of AEs |
| --- | --- | --- |
| **All** | **8 (80%)** | **55** |
| **Related to intervention** | **5 (50%)** | **18** |
| -Grade 1 (mild) | 4 (40%) | 15 |
| -Grade 2 (moderate) | 3 (30%) | 3 |
| -Grade 3-5 (severe-death) | 0 (0%) | 0 |
| -Cardiac disorder | 0 (0%) | 0 |
| -Soreness/pain | 4 (40%) | 10 |
| -Fatigue | 2 (20%) | 3 |
| -Nausea | 0 (0%) | 0 |
| -Lightheadedness | 2 (20%) | 5 |
| -Other nervous system | 0 (0%) | 0 |
| -Fall | 0 (0%) | 0 |
| -Other | 0 (0%) | 0 |
| **Possibly related or unrelated to intervention** | **8 (80%)** | **37** |
| -Grade 1 (mild) | 8 (80%) | 34 |
| -Grade 2 (moderate) | 3 (30%) | 3 |
| -Grade 3-5 (severe-death) | 0 (0%) | 0 |
| -Cardiac disorder | 0 (0%) | 0 |
| -Soreness/pain | 7 (70%) | 14 |
| -Fatigue | 0 (0%) | 0 |
| -Nausea | 0 (0%) | 0 |
| -Lightheadedness | 3 (30%) | 6 |
| -Other nervous system | 0 (0%) | 0 |
| -Fall | 5 (50%) | 11 |
| -Headache | 2 (20%) | 2 |
| -Increased knee hyperextension | 1 (10%) | 1 |
| -Joint stiffness | 1 (10%) | 1 |
| -Infection (common cold) | 1 (10%) | 1 |
| -Excision of ingrown toenail | 1 (10%) | 1 |
| -Other | 0 (0%) | 0 |

*\*There were no adverse events among control participants*

**Table 3.**
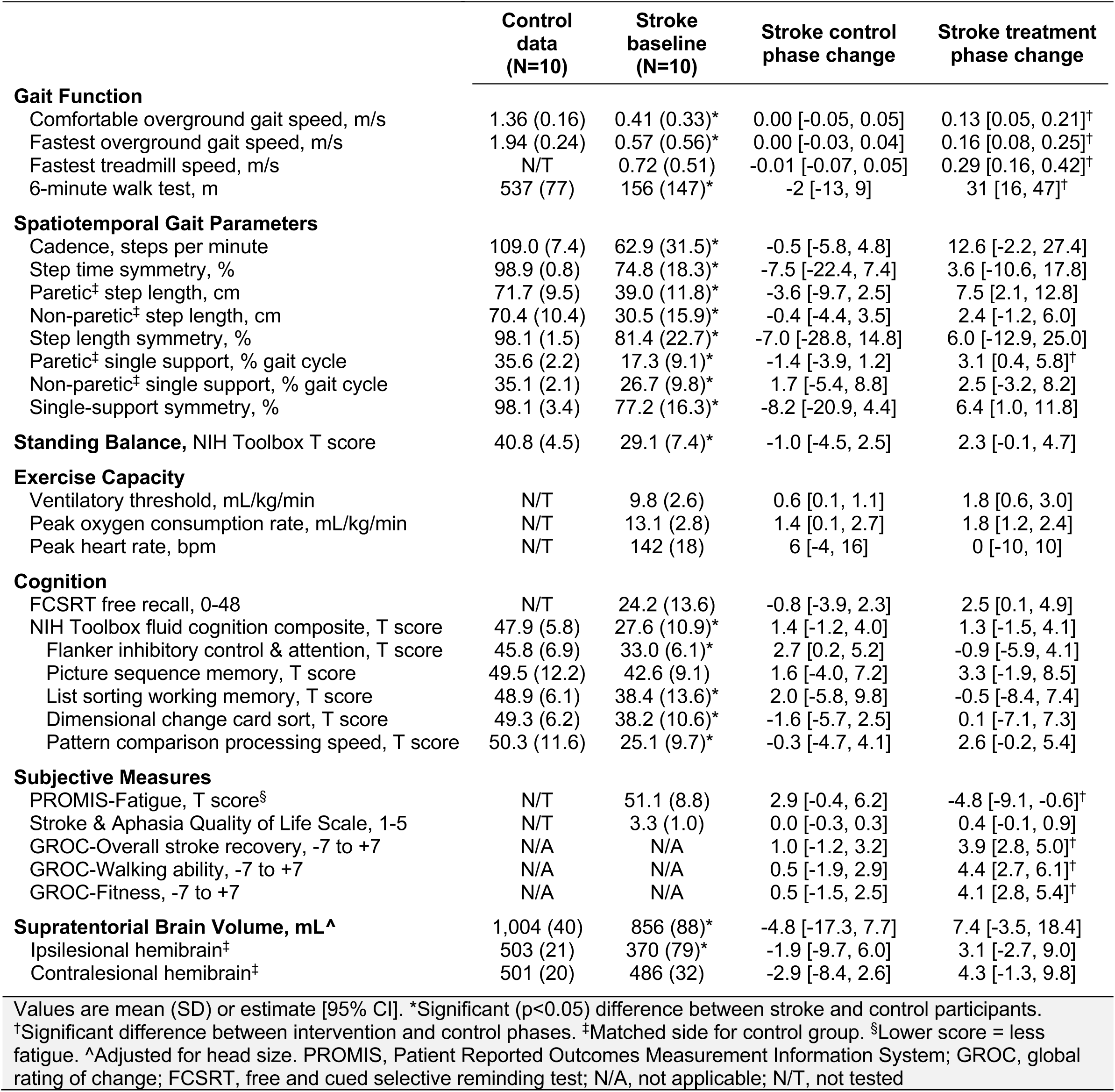
Baseline measures and outcome changes.

During the control phase, participants with stroke showed no significant change in gait function measures, spatiotemporal gait parameters, standing balance, peak heart rate, most cognitive measures, all subjective measures and brain volume (Table 2). Meanwhile, significant increases were found in ventilatory threshold, VO_2_-peak & Flanker scores. During the treatment phase, participants with stroke showed significant improvements in all gait function measures, paretic step length, paretic single support time, single-support time symmetry, ventilatory threshold, VO_2_-peak, FCSRT score, PROMIS-Fatigue score and most subjective change ratings (Table 2 & Fig 2). Compared with the control phase, treatment phase changes were significantly greater for all gait function measures, paretic single support time, PROMIS-Fatigue score, and most subjective change ratings (Table 2).

**Figure 2.**
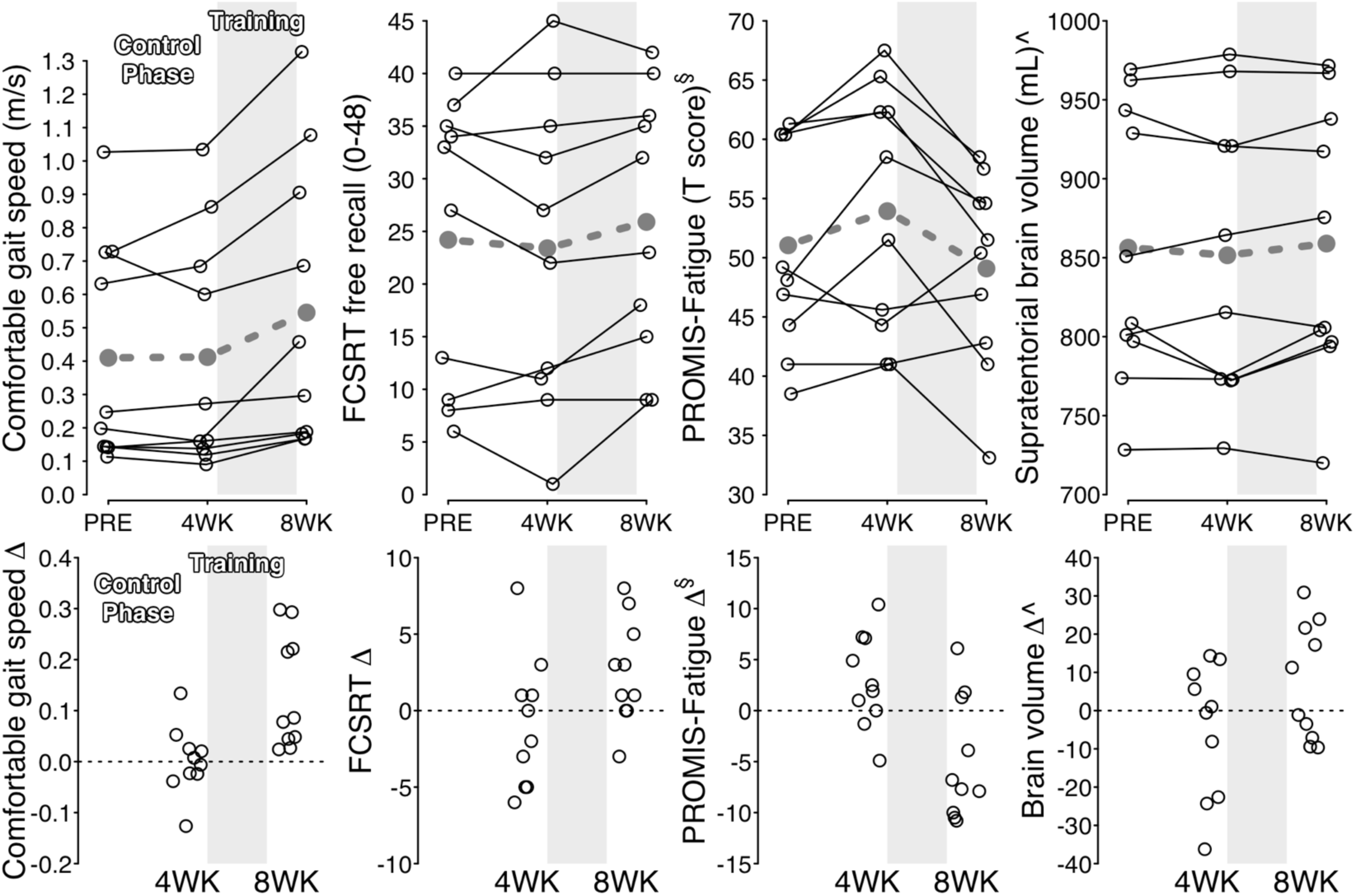
Individual changes for selected outcomes. <u>Upper panels</u>: Solid black lines show individual outcome trajectories across measurement time points. Dashed gray lines show mean estimates. <u>Lower panels</u>: Black circles show individual changes from previous time point. ^§^Lower score = less fatigue. ^Adjusted for head size. PROMIS, Patient Reported Outcomes Measurement Information System; FCSRT, free and cued selective reminding test.

When testing hypothesis 1, improvement in fastest overground gait speed during the treatment phase (+0.16 m/s [95% CI: 0.08-0.25 m/s]) averaged 61% [33-89%] of the improvement in fastest treadmill speed (+0.29 m/s [0.16-0.42 m/s]).

When testing hypothesis 2, the following outcome measures had standardized effect sizes ≥0.4 for both the treatment phase and the treatment vs. control phase comparison (Table 4): all gait function measures, cadence, paretic & non-paretic step length, paretic single-support time, single support symmetry, NIH Toolbox standing balance test, ventilatory threshold, FCRST free recall, NIH Toolbox pattern comparison processing speed test, PROMIS-Fatigue, all subjective measures and all brain volumes.

**Table 4.**
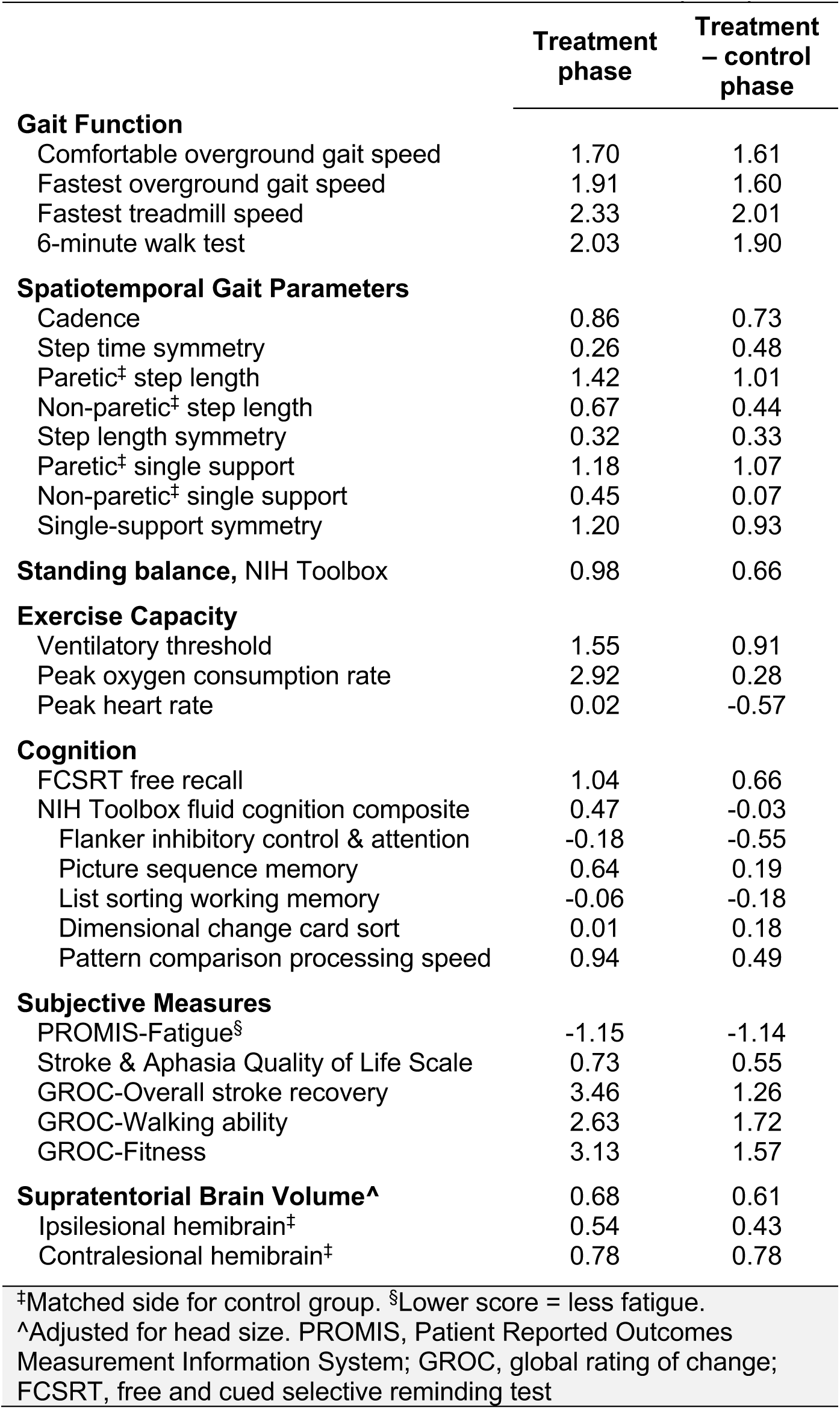
Cohen’s d standardized effect size estimates (N=10)

When testing hypothesis 3, participants with baseline comfortable gait speed ≥0.4 m/s had significantly greater absolute improvement in six-minute walk distance than participants with baseline speed <0.4 m/s (Table 5). Mean percent change in six-minute walk distance was greater in the slower baseline subgroup, but not significantly different. The slower baseline subgroup also had a significantly greater percent improvement in fastest treadmill speed. Both subgroups reported similar levels of improvement on subjective measures.

**Table 5.** Treatment phase changes by baseline comfortable gait speed subgroup.

|  | <0.4 m/s (N=6) | ≥0.4 m/s (N=4) |
| --- | --- | --- |
| **Gait Function** |  |  |
| Comfortable overground gait speed, m/s | 0.09 [-0.01, 0.18] | 0.20 [0.09, 0.32] |
| Comfortable overground gait speed, % baseline | 61.8 [11.9, 111.7] | 25.0 [-36.1, 86.1] |
| Fastest overground gait speed, m/s | 0.11 [0.01, 0.21] | 0.25 [0.13, 0.37] |
| Fastest overground gait speed, % baseline | 62.8 [14.1, 111.5] | 24.3 [-35.4, 83.9] |
| Fastest treadmill speed, m/s | 0.28 [0.10, 0.45] | 0.31 [0.10, 0.53] |
| Fastest treadmill speed, % baseline | 81.2 [46.9, 115.5] | 26.2 [-15.8, 68.2]* |
| 6-minute walk test, m | 17 [6, 28] | 53 [39, 66]* |
| 6-minute walk test, % baseline | 37.1 [23.5, 50.7] | 18.8 [2.2, 35.5] |
| **Subjective Measures** |  |  |
| PROMIS-Fatigue, T score<sup>§</sup> | -5.5 [-11.4, 0.4] | -3.8 [-11.0, 3.4] |
| Stroke & Aphasia Quality of Life Scale, 1-5 | 0.5 [-0.1, 1.2] | 0.1 [-0.7, 0.9] |
| GROC-Overall stroke recovery, -7 to +7 | 4.5 [3.1, 5.9] | 3.0 [1.3, 4.7] |
| GROC-Walking ability, -7 to +7 | 4.8 [2.5, 7.1] | 3.8 [0.9, 6.6] |
| GROC-Fitness, -7 to +7 | 3.7 [1.9, 5.4] | 4.8 [2.6, 6.9] |

Values are the mean estimate [95% CI]. \*Significant difference between gait speed subgroups.
<sup>§</sup>Lower score = less fatigue. PROMIS, Patient Reported Outcomes Measurement Information System; GROC, global rating of change

## DISCUSSION

This pilot study aimed to guide future locomotor HIIT research in chronic hemiparetic stroke by: 1) assessing overground translation of treadmill speed gains with a refined treatment protocol that included both overground and treadmill training; 2) evaluating responsiveness of different outcome measures across domains, including gait, balance, fitness, cognition, fatigue/subjective and brain volume; and 3) examining the potential for moderation by baseline gait speed. Non-stroke control participants were also included to facilitate interpretation of baseline measures. Participants with stroke showed expected differences from controls, with lower function or greater impairment in each domain tested. Consistent with our hypotheses, we found that: 1) improvement in fastest overground gait speed averaged >50% of the improvement in fastest treadmill speed (61%); 2) some outcome measures in each domain showed standardized effect sizes ≥0.4; and 3) participants with faster baseline gait speed appeared to have greater absolute improvement in 6-minute walk distance, yet similar proportional change and perceived improvement. Due to very large observed effect sizes, some variables also exhibited statistically significant improvement during the treatment phase, significant effect moderation by baseline gait speed and significant treatment vs. control phase comparisons.

Using combined treadmill and overground HIIT, the percentage of treadmill speed gains that translated into overground gait in this study was double that observed in our previous study using treadmill-only HIIT^6^ (61% vs. 30%). While comparisons across studies can be misleading (e.g. due to differences in prognosis or treatment delivery between study samples), these likely sources of confounding should have mostly worked against the current study. For example, the current sample had lower mean baseline comfortable gait speed than our previous study (0.41 m/s vs. 0.63 m/s), indicative of worse prognosis for overground gait improvement. Also, the current treatment protocol did not attempt to reduce treadmill handrail support to approximate the dynamic balance challenge of overground gait as we had done previously. Thus, the magnitude of the between-study difference and the likely direction of potential confounding suggest that overground HIIT may have been an important addition to the intervention protocol to continue in future studies. While translation was still not complete (i.e. 100%), we suspect that it may be close to the feasible limit, because the fixed balance & posture support provided by an optimally-positioned treadmill handrail cannot be entirely replicated with an assistive device during overground gait.

Our findings may also be useful for guiding selection of outcome measures in future post-stroke HIIT research and related intervention studies. It has been suggested that only interventions and outcome measures with preliminary Cohen’s d effect sizes ≥|0.4| should be advanced towards efficacy trials in neurologic rehabilitation.^25^ Based on this criterion, the results in Table 4 indicate that some outcomes in each domain (gait function, spatiotemporal gait parameters, balance, exercise capacity, cognition, subjective measures & brain volume) had sufficient responsiveness to consider including in future related studies. Large effect sizes (≥|0.8|) were observed for both the treatment phase and treatment vs. control phase comparison for all gait function measures, select gait parameters (paretic step length, paretic single support time & single support symmetry), ventilatory threshold, PROMIS-Fatigue and subjective change ratings. Moderately large effect sizes (≥|0.6|) were found for cadence, NIH Toolbox standing balance test, FCSRT free recall, and brain volume. The estimated effect sizes should be viewed with caution due to the small sample size, which makes them more prone to fluctuate with the addition of more data. However, these preliminary results may assist with prioritization of outcome measures in future studies. They also provide justification for adding cognitive, fatigue and brain volume measures to the typical motor function and fitness assessment batteries used in locomotor HIIT research.

Unexpectedly, a few outcome measures had significant changes during the no-intervention control phase, when participants were expected to be stable. This included ventilatory threshold, VO_2_peak, and the NIH Toolbox Flanker inhibitory control & attention test. While these results could plausibly be false positives due to the number of variables tested, they could also indicate learning or conditioning effects from repeated testing. This emphasizes the importance of appropriate control groups in future studies, even in chronic stroke.

The current findings also suggest that future locomotor HIIT studies should consider baseline gait speed as a potential stratification factor for randomization and/or a potential covariate in statistical models. Consistent with previous gait training research post-stroke,^21-24^ we found significantly greater absolute improvement in six-minute walk distance among participants with baseline gait speed ≥0.4 m/s vs. <0.4 m/s, despite having extremely low power to test for such interactions. While it could be argued that future studies should exclude persons with speed <0.4 m/s, we would advocate against this for several reasons. First, the 95% confidence interval for change in six-minute walk distance included the possibility of a meaningful effect even in this subgroup. Proportional changes and perceived changes in this subgroup were also comparable or better than the faster walking subgroup. In addition, the treatment protocol had a relatively short 4-week duration, and it remains unknown whether persons with baseline speed <0.4 m/s may be more responsive with a longer locomotor HIIT protocol. Finally, there are limited available interventions with evidence of efficacy for persons with severe gait impairment in chronic stroke. Thus, we would recommend continued inclusion of persons with comfortable gait speed <0.4 m/s in future studies, while incorporating baseline gait speed as a stratification factor and covariate.

The observed safety data in this study were similar to our previous research on locomotor HIIT and moderate-intensity exercise in chronic stroke.^6^ No serious AEs occurred and the frequency & type of mild/moderate AEs were consistent with expectations for exercise testing & training in a population with substantial pre-existing health conditions.^67^ When considering the current results in combination with previous studies, the safety profile of locomotor HIIT appears to be reasonable to date, but continued monitoring is needed given the relatively low numbers of participants tested and the generally unsystematic collection & reporting of AE data in the literature.

## Limitations

This study was not designed to test treatment efficacy, but rather to guide future research on locomotor HIIT post-stroke. All results should be viewed as preliminary in light of the small sample size, lack of a randomized crossover order or control group, lack of an active control intervention and lack of adjustment for multiple statistical comparisons. Another limitation was that we were unable to assess any potential outcome differences between short-interval and long-interval HIIT, since each participant performed both types in alternating sessions. This was done to increase the validity of short vs. long-interval HIIT intensity comparisons for a previous analysis.^15^ The lack of rater blinding for some outcome measures also increases risk of bias.

This was partially mitigated by using standardized protocols and automated procedures where feasible. For the subjective outcomes, participants were asked to answer as honestly as possible even if they perceived no changes or negative changes, and were told that our priority was getting accurate information to guide future research. However, we cannot rule out the possibility that some may have still reported more positive results to avoid social discomfort.

## Conclusions

Future studies of locomotor HIIT in chronic stroke should consider: 1) including both overground and treadmill training; 2) measuring cognition, fatigue and brain volume, in addition to typical gait & fitness assessment; and 3) stratifying randomization by baseline gait speed and including it as a covariate in statistical models.

## Data Availability

All data produced in the present study are available upon reasonable request to the authors

## Author Contributions

Conceptualization (PB, JV, KD, BK), data curation (PB), formal analysis (PB), funding acquisition (PB, JV, KD, BK), investigation (PB, SD, VS, ES, DW), methodology (all), project administration (PB, JV), resources (PB, JV, KD, BK, RhS, DC), software (PB), supervision (PB, JV, KD, BK), visualization (PB), writing – original draft (PB), writing – review & editing (all).

